# A deep-learning workflow to predict upper tract urothelial cancer subtypes supporting the prioritization of patients for molecular testing

**DOI:** 10.1101/2023.06.14.23291350

**Authors:** Miriam Angeloni, Thomas van Doeveren, Sebastian Lindner, Patrick Volland, Jorina Schmelmer, Sebastian Foersch, Christian Matek, Robert Stoehr, Carol I. Geppert, Hendrik Heers, Sven Wach, Helge Taubert, Danijel Sikic, Bernd Wullich, Geert J. L. H. van Leenders, Vasily Zaburdaev, Markus Eckstein, Arndt Hartmann, Joost L. Boormans, Fulvia Ferrazzi, Veronika Bahlinger

## Abstract

**Background:** Urothelial carcinoma of the bladder (UBC) comprises several molecular subtypes, which are associated with different targetable therapeutic options. However, if and how these associations extend to the rare upper tract urothelial carcinoma (UTUC) remains unclear.

**Objective:** Identifying UTUC protein-based subtypes and developing a deep-learning (DL) workflow to predict these subtypes directly from histopathological H&E slides.

**Design, Setting, and Participants:** Subtypes in a retrospective cohort of 163 invasive samples were assigned on the basis of the immunohistochemical expression of three luminal (FOXA1, GATA3, CK20) and three basal (CD44, CK5, CK14) markers. DL model building relied on a transfer-learning approach.

**Outcome Measurements and Statistical Analysis:** Classification performance was measured via repeated cross-validation, including assessment of the area under the receiver operating characteristic (AUROC). The association of the predicted subtypes with histological features, PD-L1 status, and *FGFR3* mutation was investigated.

**Results and Limitations:** Distinctive luminal and basal subtypes were identified and could be successfully predicted by the DL (AUROC 95^th^ CI: 0.62-0.99). Predictions showed morphology as well as presence of *FGFR3*-mutations and PD-L1 positivity that were consistent with the predicted subtype. Testing of the DL model on an independent cohort highlighted the importance to accommodate histological subtypes.

**Conclusions:** Our DL workflow is able to predict protein-based UTUC subtypes directly from H&E slides. Furthermore, the predicted subtypes associate with the presence of targetable genetic alterations.

**Patient Summary:** UTUC is an aggressive, yet understudied, disease. Here, we present an artificial intelligence algorithm that can predict UTUC subtypes directly from routine histopathological slides and support the identification of patients that may benefit from targeted therapy.

## Introduction

Urothelial carcinomas (UCs) are malignant epithelial neoplasms arising from the urothelial lining of the urinary tract [1]. The rare upper tract UC (UTUC) represents 5- 10% of all UCs, while the remaining are urothelial bladder cancer (UBC). UTUC is frequently associated with poor prognosis, with two-thirds of patients being diagnosed at an invasive tumor stage [2]. Owing to the histological similarity between UTUC and UBC [3, 4], and the preponderance of the latter, UTUC is an understudied disease.

Transcriptome-based subtyping allowed the stratification of muscle-invasive BC (MIBC) into different molecular subtypes [5-10], and in 2020 a consensus classification identified luminal and basal as the two distinctive subtypes [1, 11]. These subtypes offer valuable support for guiding targeted therapy options. Indeed, the luminal subtype appears associated with higher responsiveness to Fibroblast growth factor receptor 3 (*FGFR3)*-targeted therapies, and the basal subtype to immunotherapies such as programmed cell death ligand 1 (PD-L1) and programmed cell death protein 1 (PD-1) inhibitors [11, 12]. For UTUC, only few studies have so far investigated its genomic and transcriptomics landscape [13-15]. Additionally, no consensus subtypes have been identified yet [3, 16].

Assessment of molecular subtypes via high-throughput sequencing is neither ubiquitously available, nor cost-effective. Thus, alternative subtyping approaches should be considered. In MIBC, studies showed a substantial overlap between molecular and protein-based subtypes [17, 18], identifiable via more widespread, routinely applicable immunohistochemistry (IHC) analyses. Moreover, AI-based approaches have recently emerged as novel research tools able to provide automated and accurate pathological diagnoses, leveraging the information residing into whole-slide images (WSIs) [19]. Here, we comprehensively characterize UTUC protein-based subtypes and propose a deep learning (DL) approach that can predict them relying on routine digitized H&E slides.

## Patients and methods

### Patient cohorts

The German cohort comprised formalin-fixed paraffin embedded material of N = 163 retrospectively analyzed UTUC patients diagnosed between 1995 and 2012 at University Hospital Erlangen-Nürnberg (Erlangen, Germany) and University Hospital Gießen and Marburg (Marburg, Germany). The Dutch cohort comprised N = 55 UTUC samples diagnosed between 2017 and 2020 as part of a multicenter, phase II, prospective trial conducted at University Medical Center Rotterdam (Rotterdam, The Netherlands) [20]. For both cohorts one whole slide image (WSI) per patient was used and all samples were invasive (i.e., with tumor stage ≥ pT1). All cases were systematically reviewed by two uropathologists (V.B., A.H.) according to the tumor, node, metastasis (TNM) classification (2017) and the classification of genitourinary tumors [21]. Clinico-pathological characteristics of the two cohorts are summarized in Table 1. The uropathologists also evaluated slides in terms of histological subtype, infiltration type, tumor type, and stroma content. Approval for this study was obtained from the Ethics Committee of the Friedrich-Alexander University Erlangen-Nürnberg (No. 329_16B) and the Erasmus Medical Centre Rotterdam (METC 2017-227 NL60919.078.17). All patients gave informed consent. The study was carried out in accordance with the Declaration of Helsinki and to the principles of Good Clinical Practice (GCP).

**Table 1.** Clinico-pathological variables characterizing the German and the Dutch cohorts. Estimates are given as median (minimum, maximum) or frequency (percentage) with respect to the total number of analyzed samples (N = 163 for the German cohort and N = 55 for the Dutch cohort). A hyphen (-) is used to indicate information/variable’s value not available within a given cohort.

| Clinico-pathological variable | German cohort | Dutch cohort |
| --- | --- | --- |
| *Age at diagnosis (yrs), median (min-max)* | 73 (47-94) | 71 (52-85) |
| *Gender, n (%)* |  |  |
| Female | 52 (31.9) | 21 (38.2) |
| Male | 111 (68.1) | 34 (61.8) |
| *Histological Grade (WHO 1973), n (%)* |  |  |
| G1 | 0 (0%) | 0 (0%) |
| G2 | 52 (31.9) | 12 (21.8) |
| G3 | 111 (68.1) | 36 (65.5) |
| Missing | 0 (0%) | 7 (12.7) |
| *Histological Grade (WHO 2004), n (%)* | - |  |
| high |  | 54 (98.2) |
| low |  | 0 (0%) |
| Missing |  | 1 (1.8) |
| *Histological Grade (WHO 2016), n (%)* |  | - |
| high | 142 (87.1) |  |
| low | 21 (12.9) |  |
| *Primary Tumor, n (%)* |  |  |
| pT1 | 33 (20.2) | 17 (30.9) |
| pT2 | 28 (17.2) | 13 (23.6) |
| pT3 | 81 (49.7) | 25 (45.5) |
| pT4 | 21 (12.9) | 0 (0%) |
| *Regional Lymph Nodes, n (%)* |  |  |
| pN0 | 54 (33.1) | 22 (40%) |
| pN1 | 16 (9.8) | 3 (5.5%) |
| pN2 | 14 (8.6) | 2 (3.6%) |
| Missing | 79 (48.5) | 28 (50.9%) |
| *Distant Metastasis, n (%)* |  |  |
| cM0 | 79 (48.5) | 54 (98.2%) |
| cM1 | 8 (4.9) | 0 (0%) |
| Missing | 76 (46.6) | 1 (1.8%) |

### Clustering-based subtype assignment and statistical analyses

For protein-based subtyping, four tumor cores were punched for each patient and stained with three luminal (CK20, FOXA1, and GATA3) and three basal (CD44, CK14, and CK5) markers at the Institute of Pathology, University Hospital Erlangen, using tissue microarrays (TMAs). Expression was quantified using the histoscore (H-Score). The same marker panel was used for IHC evaluations at the whole-slide level on selected cases. PD-L1 expression was assessed on TMAs using both immunoscore (IC) and combined positive score (CPS) [22], while *FGFR3* mutational status was assessed using the SNaPshot method [23].

Hierarchical clustering and statistical analyses were performed within the R environment v.4.0.3 [24]. Association between categorical variables was assessed using Fisher’s exact test. To compare the distribution of continuous variables, the Wilcoxon rank-sum test for independent samples (two groups), the Kruskal-Wallis test (more than two), or the one-tailed Wilcoxon signed-rank test (paired samples) were used. Analyses of overall survival (OS) and disease-specific survival (DSS) were performed using the Kaplan-Meier (K-M) estimator and statistical differences assessed through the log-rank test. P-values < 0.05 were considered statistically significant.

### WSIs annotation and pre-processing

Slides belonging to the two cohorts were digitized in the respective pathology centers using a Panoramic P250 scanner (3DHistech, Budapest, Hungary) at different resolution levels. For each WSI, tumor tissue was manually annotated in QuPath (v.0.2.3) by a trained observer (M.A.) under expert supervision (V.B.) (Supplementary Figure 1). An automated Python-based pipeline (https://github.com/MiriamAng/TilGenPro) was implemented to tessellate the identified tumor areas into non-overlapping tiles of 512×512 pixel edge length, perform quality filtering, and stain-normalization (Supplementary Figure 2).

### Deep-learning algorithm and its validation

Our DL framework relied on a transfer-learning approach by fine-tuning a ResNet50 [25] initialized with weights pre-trained on the ImageNet database [26]. To account for class imbalance, the number of tiles belonging to each class within the training set was equalized. A repeated three-fold cross-validation was used to estimate the model’s generalization accuracy and error. Here, to ensure independence between training and validation folds, random splitting was performed at patient level (Supplementary Figure 3). DL model’s prediction for single tiles of the validation set were averaged to obtain a WSI-level subtype prediction. For each repetition, the area under the receiver operating curve (AUROC), accuracy, precision, recall, and F1-Score were assessed as mean and 95% confidence interval (CI) across the three validation folds relying on Student’s t-distribution. Confusion matrices for a given repetition were obtained by concatenating the predictions for the three validation folds. The predicted class in the independent test cohort was taken equal to the one with the highest average prediction value across those obtained using the three trained models.

Further details for the methods are in the Supplementary Material.

## Results

### Hierarchical clustering of protein marker expression identifies luminal, basal, and indifferent UTUC subtypes

To characterize protein-based UTUC subtypes, the expression of three basal (CK5, CK14, CD44) and three luminal (CK20, FOXA1, GATA3) markers utilized in published UBC studies [17, 18] was assessed in a cohort of 163 invasive samples, referred to as ‘German cohort’. Hierarchical clustering of marker expression identified: a ‘luminal’ cluster (80 samples, 49%), a ‘basal’ cluster (42 samples, 26%), and an ‘indifferent’ cluster (41 samples, 25%) with low expression of both basal and luminal markers (Figure 1A). The basal samples exhibited shorter overall survival (OS) and disease-specific survival (DSS) than the luminal and indifferent samples (Figure 1B). Tumor stages differed across the three subtypes (p = 0.01), with almost half of pT4 samples in the basal group (Supplementary Table 1). Both infiltration (p = 0.02) and tumor type (p = 0.02) differed between basal and indifferent cases (Figure 1 C-D). In contrast, no significant differences were found when comparing luminal and indifferent subtypes. Indeed, basal samples showed a clear prevalence of diffusely infiltrative and non-papillary tumors, while the other two subtypes showed a higher proportion of pushing and papillary tumors. Also, luminal and indifferent subtypes were characterized by a comparable proportion of fibrotic and leiomyomatous samples, while the basal subtype had a prevalence of fibrotic stroma (Figure 1E).

**Figure 1.**
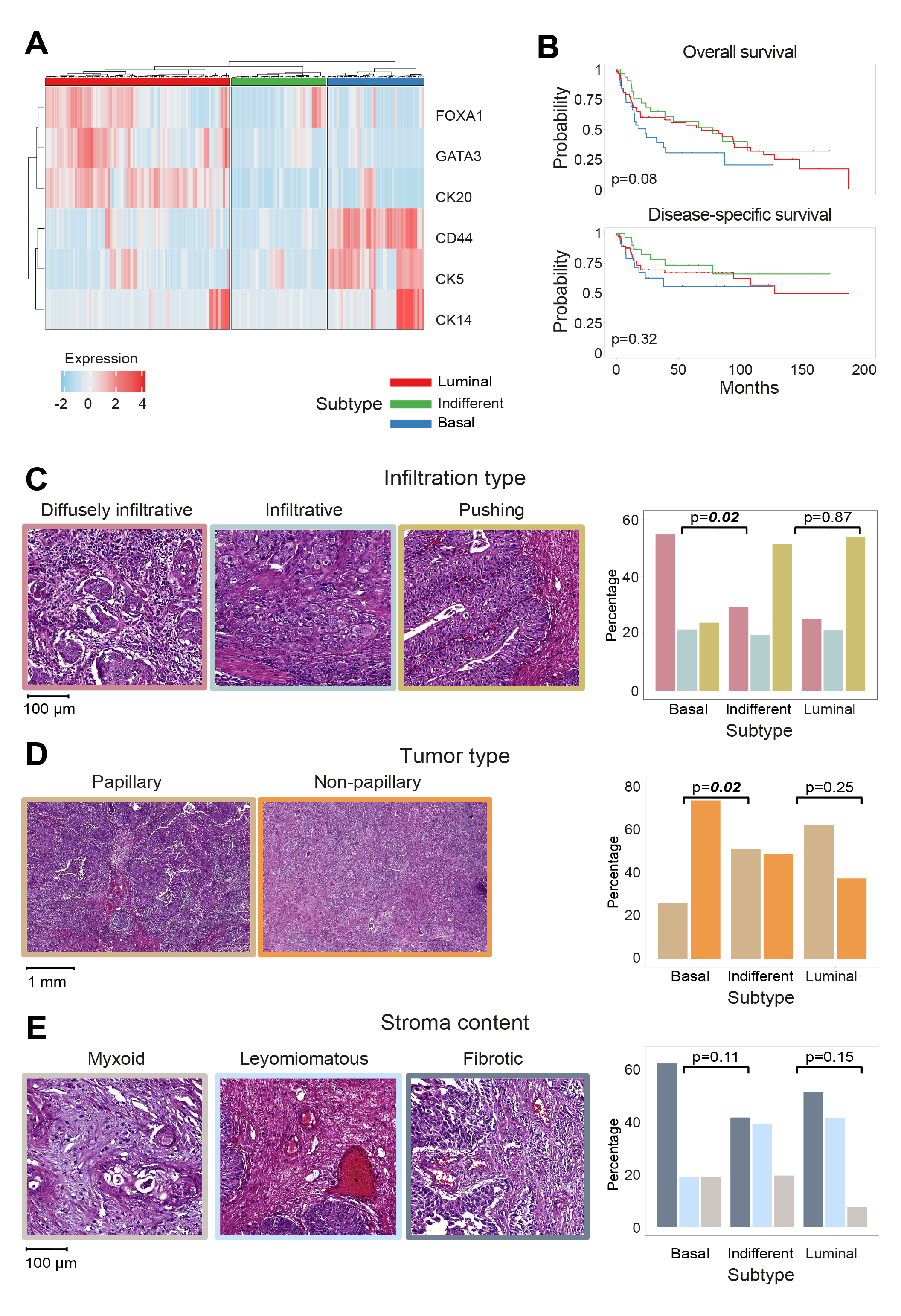
Protein-based UTUC subtypes and their histopathological characterization. (A) Heatmap visualization of the hierarchical clustering analysis performed on the expression of the three basal (CD44, CK5, CK14) and three luminal (FOXA1, GATA3, CK20) markers in our UTUC German cohort (N = 163 invasive samples). Heatmap colors represent markers expression (standardized H-Score; red: high expression, blue: low expression). The color ribbon at the top of the heatmap indicates the three protein-based subtypes: luminal (red), indifferent (green), basal (blue). (B) Kaplan-Meier overall survival (top) and disease-specific survival (bottom) curves for the identified subtypes. P-values from log-rank test. (C-E) Characterization of the identified subtypes in terms of infiltration type (C), tumor type (D) and stroma content (E). On the left representative histology images and on the right bar plot distributions. The histology images are framed in the respective color used in the bar plot to the right. P-values from Fisher’s exact tests (p-values < 0.05 in italic bold).

Collectively, hierarchical clustering based on the IHC expression of six well-characterized UBC markers identified three distinctive protein-based UTUC subtypes, with high histological similarity of the indifferent to the luminal subtype.

### Deep-learning successfully predicts luminal and basal subtypes and identifies candidate heterogeneous slides

We hypothesized that an AI approach could predict the identified subtypes from routinely available H&E slides. To test this hypothesis, 100,178 luminal, 66,770 basal and 57,874 indifferent tumor patches from the 163 annotated tumor areas of the German cohort were used to train and validate a DL-based classifier in a three-time repeated three-fold cross-validation setting (Supplementary Table 2; Supplementary Figure 4). While the model correctly predicted most basal (AUROC = 0.77; 95% CI, 0.67-0.86; repetition three) and luminal (AUROC = 0.71; 95% CI, 0.44-0.99; repetition two) samples, more than 55% of indifferent samples were predicted luminal. This difficulty of the DL model in predicting the indifferent subtype was consistent with the observed histomorphological similarity between luminal and indifferent subtypes.

Therefore, we decided to train the model focusing on the most distinctive luminal and basal subtypes, again relying on a repeated cross-validation setting (Figure 2A). Our classifier achieved a mean AUROC of at least 0.8 (AUROC = 0.83; 95% CI, 0.67- 0.99; repetition one) and mean accuracy of ≥ 0.75 (mean accuracy = 0.79; 95% CI, 0.75-0.84; repetition two) (Supplementary Table 3). For further analyses, we focused on the results of repetition 2, as it showed the best accuracy and most consistent performance metrics across the three hold-out folds (Supplementary Table 3). First, the so called “high-confidence” slides were identified, i.e., those slides predicted luminal or basal with a score of at least 0.7, irrespective of their true subtype (Supplementary Table 4). The true positive rates in the high-confidence luminal and basal slides were respectively 86% (50/58) and 87.56% (14/16). Visual inspection of tile-level prediction maps of the top high-confidence slides confirmed the pathological description of these subtypes at histological level, i.e., dense nuclei with small stroma bridges as distinctive feature of the luminal subtype (Figure 3A) and dense stroma and keratinization for the basal subtype (Figure 3B) [1]. In the top scoring luminal slide, 99.9% of tiles were predicted luminal. In the top basal, 90% of tiles were predicted as basal. These predictions were confirmed by whole-slide level staining with the six luminal and basal markers (Supplementary Figure 5 A-B), thus suggesting that our DL model can successfully distinguish between luminal and basal subtypes.

**Figure 2.**
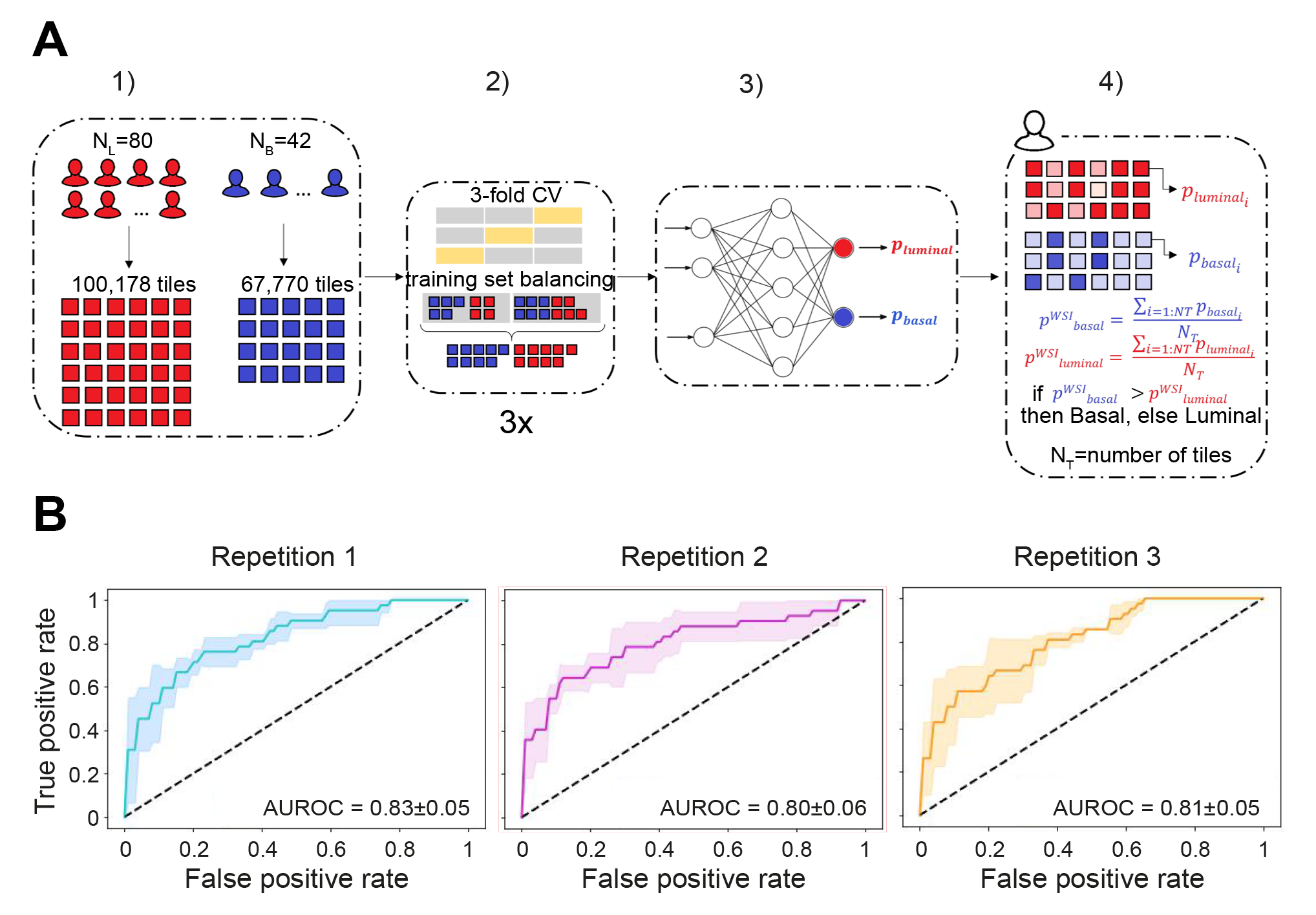
Deep-learning based prediction of luminal and basal protein-based subtypes. (A) Steps of the deep learning framework: (1) Starting from 80 luminal and 42 basal WSIs, a library made up of 100,178 luminal (red) and 66,770 basal (blue) stain-normalized tiles is generated using an automated, custom-developed, pre-processing pipeline. (2) The tiles library is used for training the network using three-fold cross validation (grey: training folds, yellow: validation fold). Tiles of the trained set are balanced between the two classes. The cross-validation is repeated three times. (3) For each training/validation set combination, a deep-learning model is trained using a transfer-learning approach. For each tile, the model outputs a prediction value for the luminal (p_luminal_) and for the basal (p_basal_) class (4). WSI-level predictions for the luminal (p^WSI^_luminal_) and basal (p^WSI^_basal_) subtypes are calculated by averaging the tile-level predictions for each class. The subtype associated with the highest prediction is assigned to the entire slide (4). In the schematization color intensity is proportional to the prediction score. (B) AUROC for the three repetitions (with basal type as positive class). The mean AUROC ± standard deviation across folds is reported for each repetition. AUROC: area under the receiver operating characteristic; N_L_: number of luminal slides; N_B_: number of basal slides.

**Figure 3.**
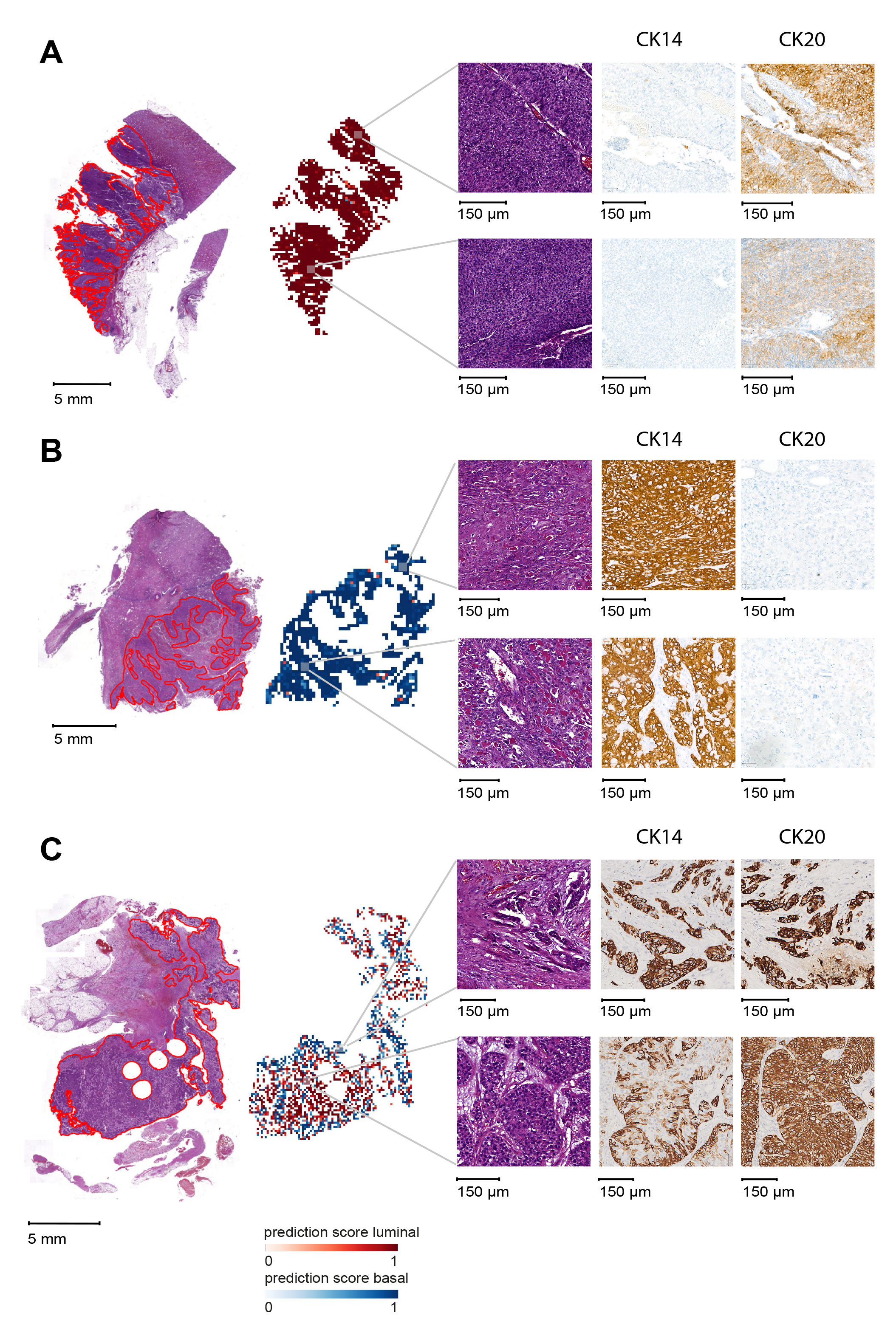
Whole-slide IHC-validation of deep-learning predictions. (A) Slide predicted with the highest p^WSI^_luminal_; (B) Slide with the highest p^WSI^_basal_; (C) Candidate heterogeneous slide. For all three slides, from left to right: digitized whole-slide with annotated tumor areas (red); tile-level prediction map (red: luminal, blue: basal; intensity dependent on prediction score); selected areas and corresponding whole-slide IHC-validation using CK20 as representative luminal marker and CK14 as representative basal marker. Whole-slide IHC-validation with all the six luminal/basal markers is provided in Supplementary Figure 5.

Next, we focused on 22 ‘low-confidence’ predictions with prediction scores between 0.4 and 0.6 (Supplementary Table 5). These slides showed no significant difference in the distribution of luminal and basal marker expression (p = 0.43). Tile-level prediction maps allowed categorizing these slides into: ‘heterogeneous slides’ with and slides without distinguishable clusters of predicted luminal and basal tiles. Visual inspection of a selected candidate heterogeneous slide supported the predictions, with basal and luminal tiles showing the characteristic histological features (Figure 3C). Furthermore, whole-slide IHC validation showed positivity of the entire tumor area for the three luminal markers and the basal marker CK14 (Supplementary Figure 5C). Notably, in the luminal-predicted area, the CK14 basal marker appeared expressed only in the outer cellular layer, while in the basal-predicted area it appeared in all cell layers (Figure 3C). Taken together, the DL model was able to identify candidate heterogenous slides showing co-presence of luminal and basal areas.

### High-confidence predicted slides show the expected histology features and significantly associate with PD-L1 and *FGFR3* status

To further characterize the high-confidence predictions, we examined also for them marker expression and morphological characteristics. High-confidence predicted luminal and basal slides showed higher expression of luminal (p = 6.62*10^-9^) or basal markers (p = 0.002), respectively (Figure 4A).

**Figure 4.**
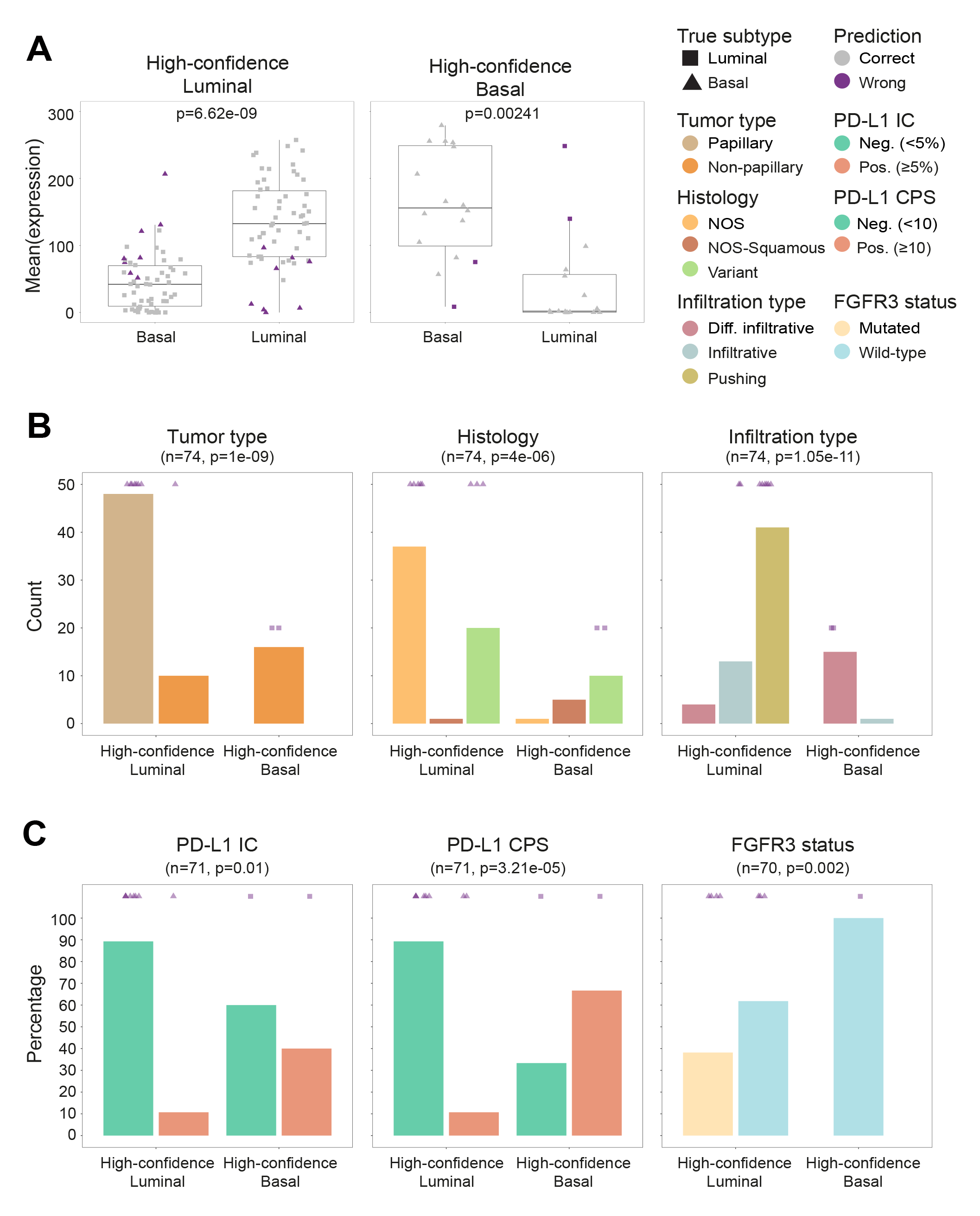
Characterization of the high-confidence predicted luminal and basal slides. (A) Boxplot distributions of the mean expression values for the basal (CK14, CK5, CD44) and luminal (CK20, FOXA1, GATA3) markers in the high-confidence predicted luminal (left) and basal (right) slides. Shape represents the true slide label (triangle: basal; square: luminal); color represents the prediction accuracy (grey: correct predictions; violet: incorrect predictions). P-values from one-tailed Wilcoxon signed-rank test. (B) Characterization of the high-confidence predicted slides in terms of tumor type (left), histology (middle), and infiltration type (right). An overview of the morphological features for the wrongly predicted slides is provided through the colored symbols above the bars. (C) Characterization of the high-confidence predicted slides in terms of clinical-relevant biomarkers: PD-L1 status (measured as CPS = combined positive score; IC = immunoscore) and *FGFR3* mutational status. An overview of PD-L1 and *FGFR3* status of the wrongly predicted slides is provided through the colored symbols above the bars. n: number of samples; p-values from one-tailed Fisher’s exact test.

High-confidence luminal predictions were mainly characterized by papillary tumors, with not otherwise specified (NOS) or histological subtype, and with pushing infiltration type. Instead, high-confidence basal predictions were mainly non-papillary tumors either squamous or with a histological subtype component and diffusely infiltrating (Figure 4B). Of note, five out of the eight wrongly predicted luminal slides were pushing papillary tumors with NOS histology, whereas both wrongly predicted basal slides were diffusely infiltrating, non-papillary tumors with variant histology. These results show that high-confidence predictions showed morphologic features consistent with the predicted subtype.

Next, we investigated the association of high-confidence slides with clinically-relevant biomarkers (Figure 4C). The proportion of PD-L1-positive samples was higher in high-confidence predicted basal slides, both using IC (p = 0.01) and CPS (p < 0.001). Vice versa, the proportion of *FGFR3*-mutated samples was higher in high-confidence predicted luminal slides (p-value=0.002). Interestingly, one wrongly predicted basal slide was actually PD-L1-positive, whereas four wrongly predicted luminal slides were *FGFR3* mutated.

### External validation of the deep-learning model highlights the importance of subtype histology

To test the generalization ability of the DL model, an external cohort of 55 invasive UTUC patients, referred to as the ‘Dutch cohort’, was used. Hierarchical clustering identified also in this cohort luminal (31 samples, 56%), basal (4 samples, 7%), and indifferent (20 samples, 36%) subtypes, with expression profiles matching those in the German cohort (Supplementary Figure 6A).

The DL model correctly classified all luminal samples, with an average prediction score of 0.89, but not the four basal samples (Supplementary Figure 6B). Three of these samples showed histological subtypes with glandular, squamous and sarcomatoid features; one sample was instead predominantly papillary with NOS histology. However, in this fourth sample, basal features could be observed at the invasion front, where two out of four TMA cores were punched. Interestingly, the tile-level prediction map highlighted a small tumor area predicted basal in correspondence to the invasion front, whereas the remaining papillary area was predicted luminal (Supplementary Figure 6C). Thus, the difficulties in predicting the basal samples might be due to histological subtype.

## Discussion

We identified three protein-based UTUC subtypes, which partially resembled UBC protein-based subtypes [17, 18]. The indifferent subtype, similarly to UBC double negative, presented low expression of both luminal and basal IHC markers, and would require further validation using more comprehensive methods such as RNA sequencing.

Our characterization in terms of infiltration type, tumor type, and stroma content, shed light into the histological similarity between the indifferent and luminal subtypes. This similarity was reflected in the performance of a three-class DL model, which predicted a large number of indifferent samples as luminal. Instead, we showed that a DL model could successfully discriminate between the most distinctive luminal and basal subtypes relying on routinely available H&E slides. Furthermore, the DL model identified candidate heterogeneous slides for which whole-slide IHC validation confirmed the co-presence of luminal and basal areas closely matching the tile-level predictions.

At histopathology level, invasive UCs present different morphological appearances [1]. As observed in Weyerer et al., MIBC histological subtypes are strong indicators of mRNA-/protein-based subtypes. Notably, squamous differentiation, characterized by intercellular bridges or keratinization, only occurs in the basal subtype, whereas conventional UCs (NOS) are mainly associated with the luminal subtype [17]. The high-confidence predictions by our model, even including those slides where the predicted subtype did not match the expression-based subtype, showed histological features consistent with the predicted subtype. Moreover, high-confidence predicted luminal and basal slides were significantly associated with *FGFR3* and PD-L1, respectively. Because of the implementation of anti-PD-L1 and PD-1 therapies, and specific *FGFR3* inhibitors [21, 27] in UCs, histopathology labs have been facing increased requests for PD-L1 assessment and *FGFR3* genetic alteration testing. Our DL model thus offers a valuable support to pathologists for the prioritization of *FGFR3*/PD-L1 testing in UTUC patients. This would also contribute to extending patients’ access to targeted therapies and improve their management and care in the clinical practice.

Several challenges were encountered during our study. Hierarchical clustering in the independent Dutch cohort highlighted the same biological tendency observed in the German cohort, which is even more remarkable considering the prospective nature of the former. This strongly supported the existence of three distinct UTUC subtypes. However, the cluster membership of single samples might vary with varying samples being clustered. This uncertainty in the training sample labels might negatively affect the DL model performance. An additional level of uncertainty in training labels was due to the assessment of marker expression in selected TMA cores. It is common practice to stain several tumors at once, while also taking into account tumor heterogeneity. However, expression in TMA cores might not be representative of whole-slide level expression, as clearly seen for the predominantly papillary case in the Dutch cohort with a basal-like morphology at the invasion front. RNA-sequencing analyses might offer a more robust subtype assignment, thanks to genome-wide profiling, yet might still fail to correctly characterize heterogeneous samples. Another challenge was linked to histological subtypes. Given the rarity of UTUC, we had decided not to exclude them, as instead done in previous UBC studies [18]. However, as the results on the Dutch cohort showed, this might have impaired model performance. This issue could be mitigated by the future availability of an even more extensive UTUC cohort.

Collectively, our results show that the most distinctive protein-based UTUC subtypes can be predicted directly from H&E slides and are associated with the presence of targetable genetic alterations. Thus, our study lays the foundation for an AI-based tool to support UTUC diagnosis and extend patients’ access to targeted treatments.

## Data Availability

All data produced in the present study are available upon reasonable request to the authors.

https://github.com/MiriamAng/TilGenPro

## Supplementary Figure Legends

**Supplementary Figure 1.**
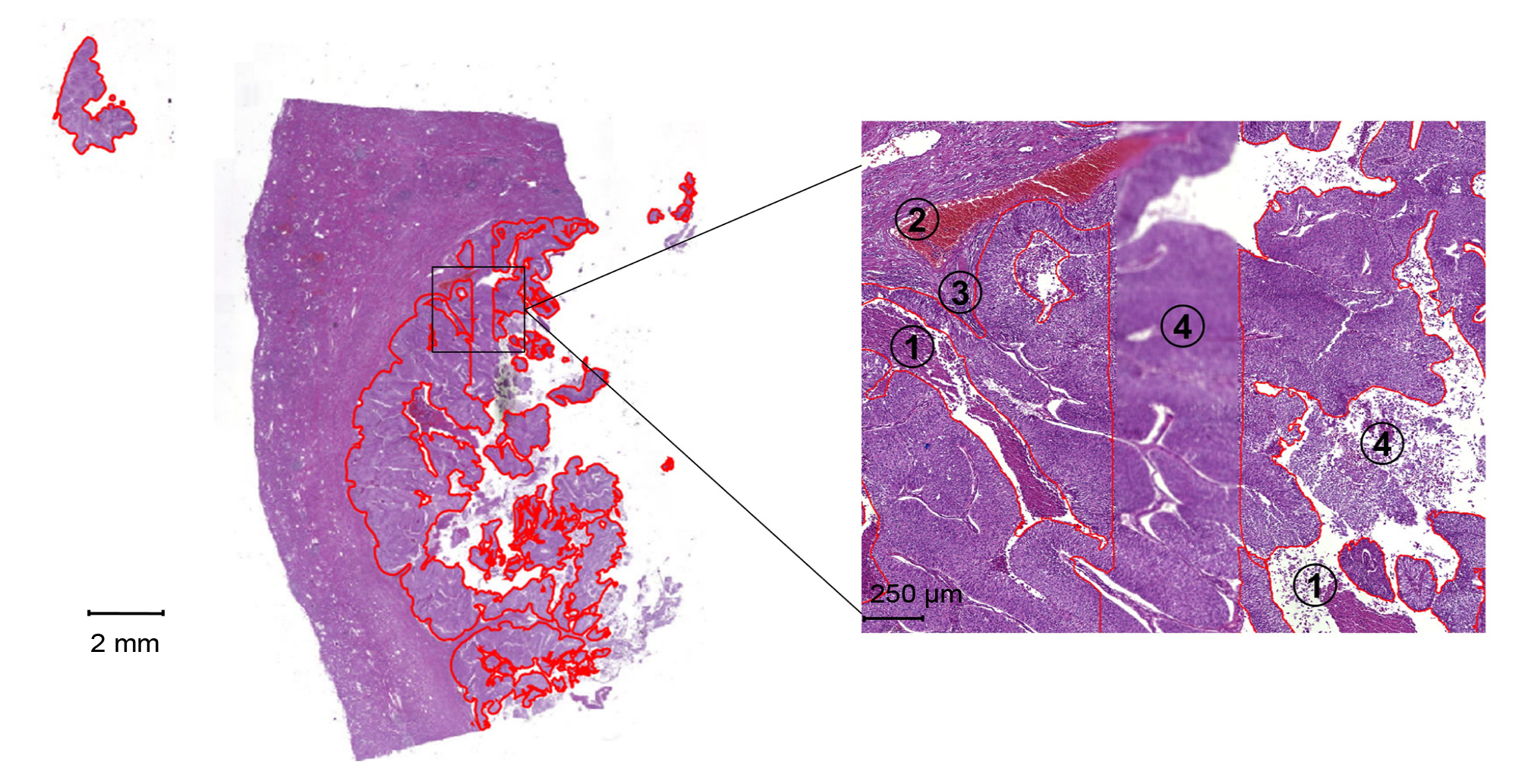
Example of tumor annotation. (left) Overview of a digitized whole slide image (WSI) with tumor tissue annotation borders in red; (right) zoomed-in tumor area representative of the employed annotation criteria: necrosis (1), bleeding/blood vessels (2), peri-tumoral lymphocytes (3) and scanning artifacts such as blurring and poor fixation (4) were excluded from the annotated area.

**Supplementary Figure 2.**
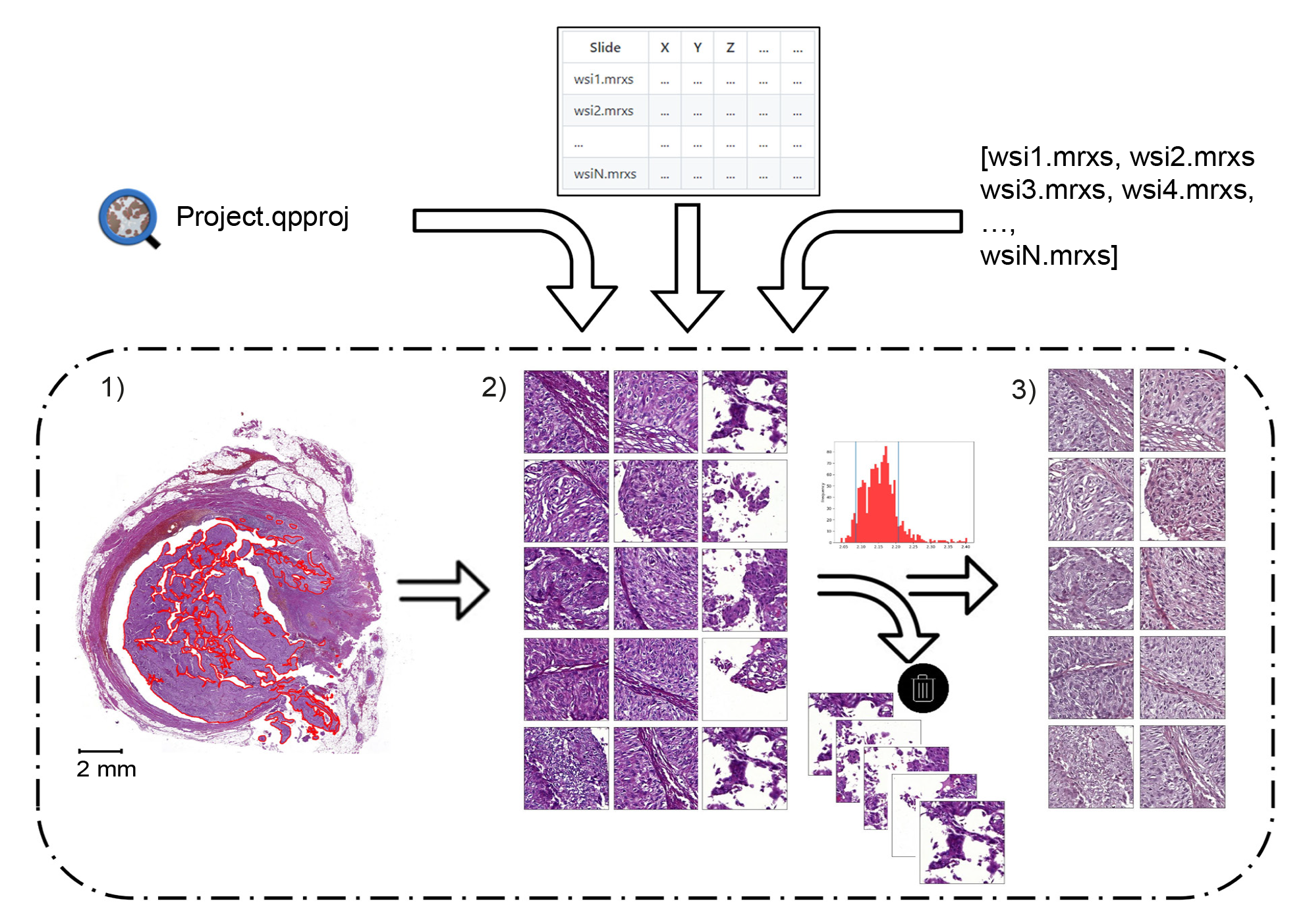
Overview of the WSIs pre-processing workflow. Graphical representation of the main steps performed by the automated Python-based pre-processing pipeline. A QuPath project (Project.qpproj) or a set of WSIs to process (provided via a csv file, or specified individually as a list of one or more WSI names) can be used as input for the pipeline. The pipeline performs the following steps: (1) The annotated tumor area (red) within each slide is tessellated into smaller tiles; (2) that the tiles undergo a quality-filtering step based on lower/upper thresholds identified on the log10-transformed median intensity values distribution; (3) tiles are stain-normalized according to the Macenko method.

**Supplementary Figure 3.**
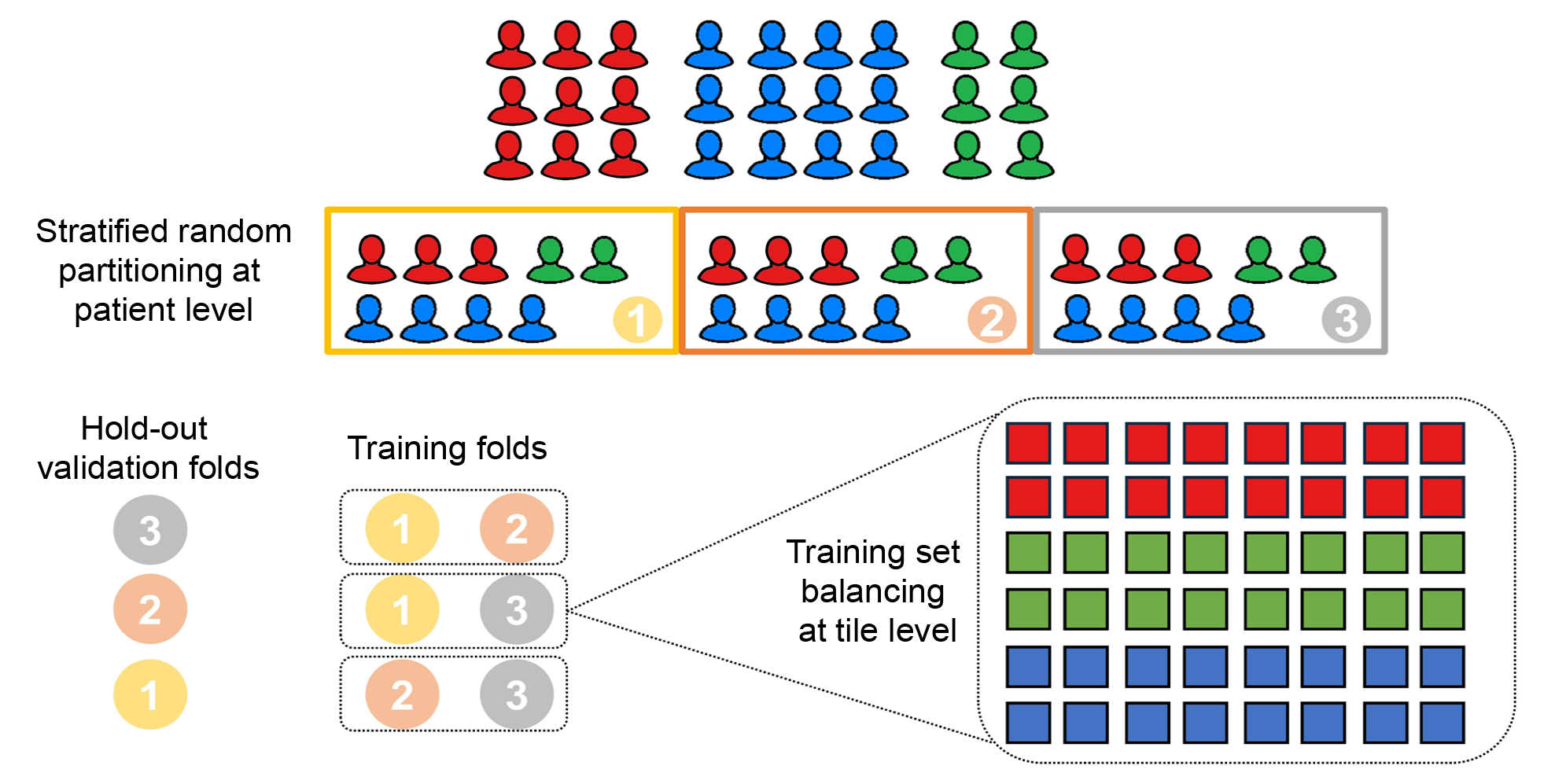
Overview of the training set generation procedure in a three-fold cross-validation setting. In the example, each icon represents a patient (in our case corresponding to a slide, as only one slide per patient was used) of the analyzed cohort and the three colors (red, blue and green) represent three different classes of patients. The patients’ cohort is randomly split in three stratified partitions, i.e., with the same class distribution as in the whole cohort. Two partitions out of three are used, in turn, to train the deep-learning model while the correspondent hold-out folds are used for validation. Tiles belonging to the two training partitions are pooled together and class. In the example tiles are represented by the square symbol and inherit the same color (i.e., class) of the parent patient.

**Supplementary Figure 4.**
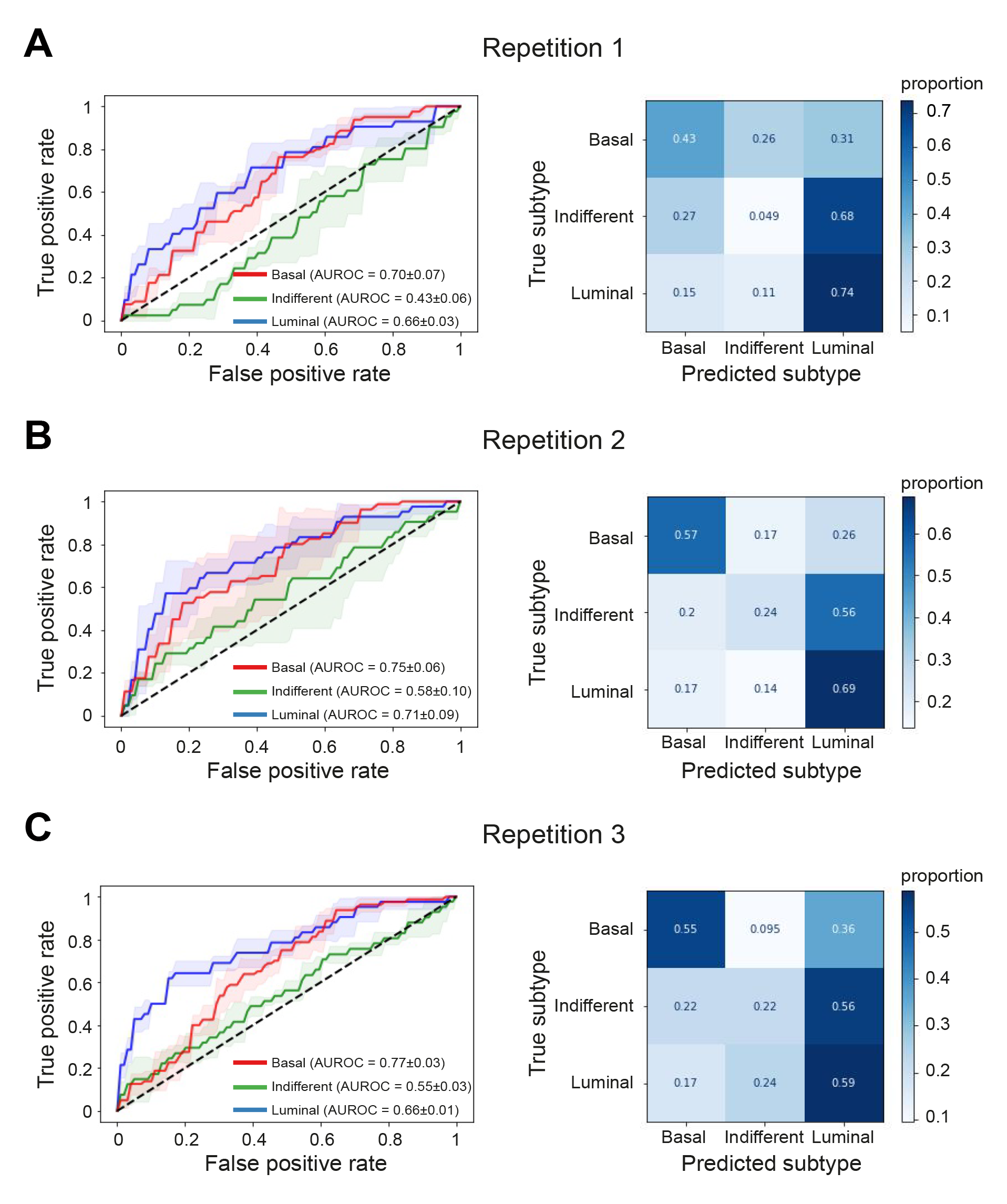
Deep-learning model performance in the prediction of the luminal, basal, and indifferent subtypes. AUROC curves and confusion matrices for (A) repetition 1, (B) repetition 2 and (C) repetition 3 of the three-fold cross-validation. AUROC curves are shown for each subtype (blue: basal, green: indifferent, red: luminal). The mean AUROC ± standard deviation (sd) is reported for each repetition. Confusion matrices are normalized over the true class (row). AUROC: area under the receiver operating characteristic.

**Supplementary Figure 5.**
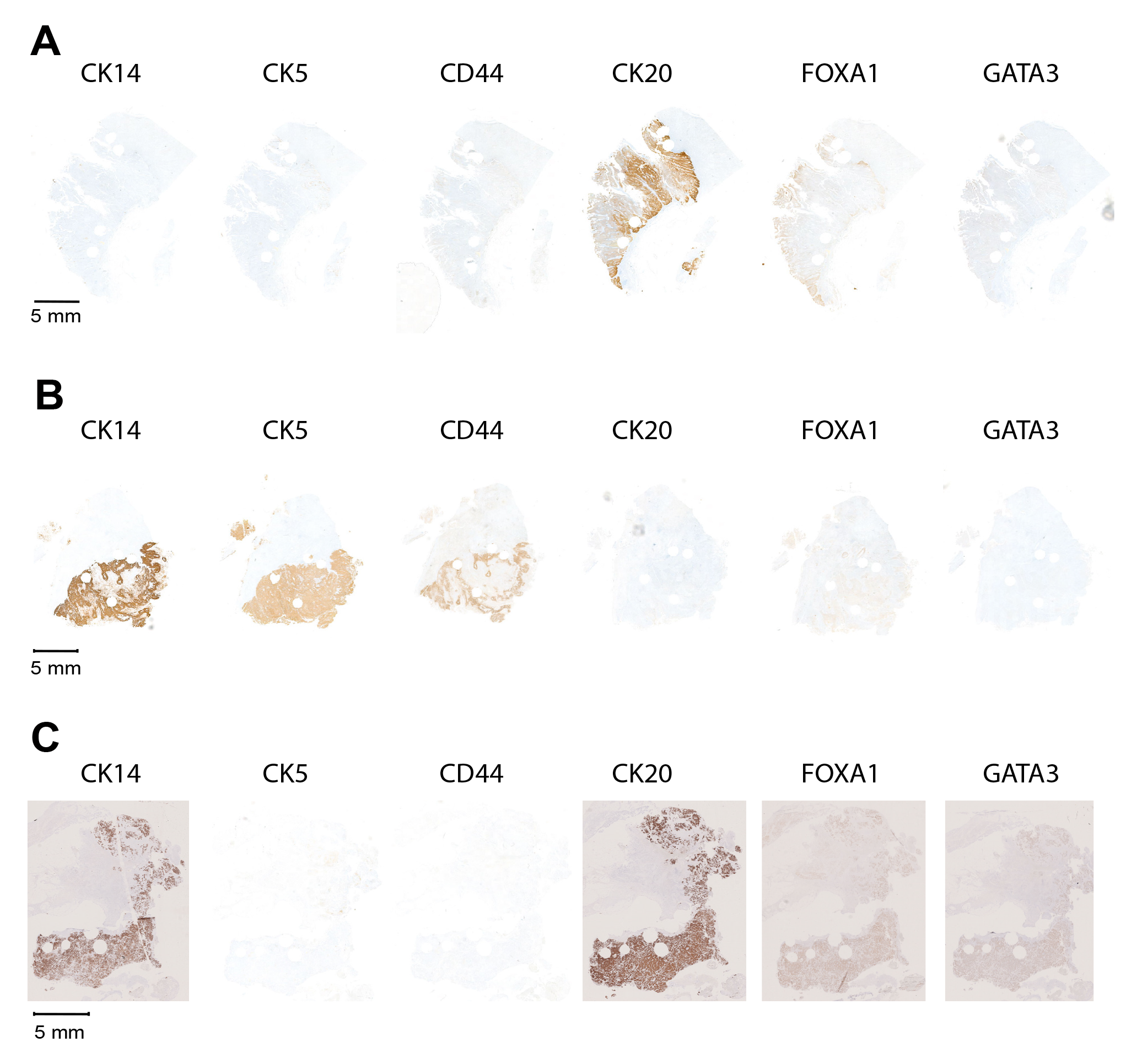
Whole-slide IHC-validation with the entire markers panel of the samples shown in Figure 3. Whole-slide IHC-validation with the three basal (CK14, CK5, CD44) and three luminal (CK20, FOXA1, GATA3) markers for (A) the top high-confidence predicted luminal slide, (B) the top high-confidence predicted basal slide, and (C) a candidate heterogeneous slide.

**Supplementary Figure 6.**
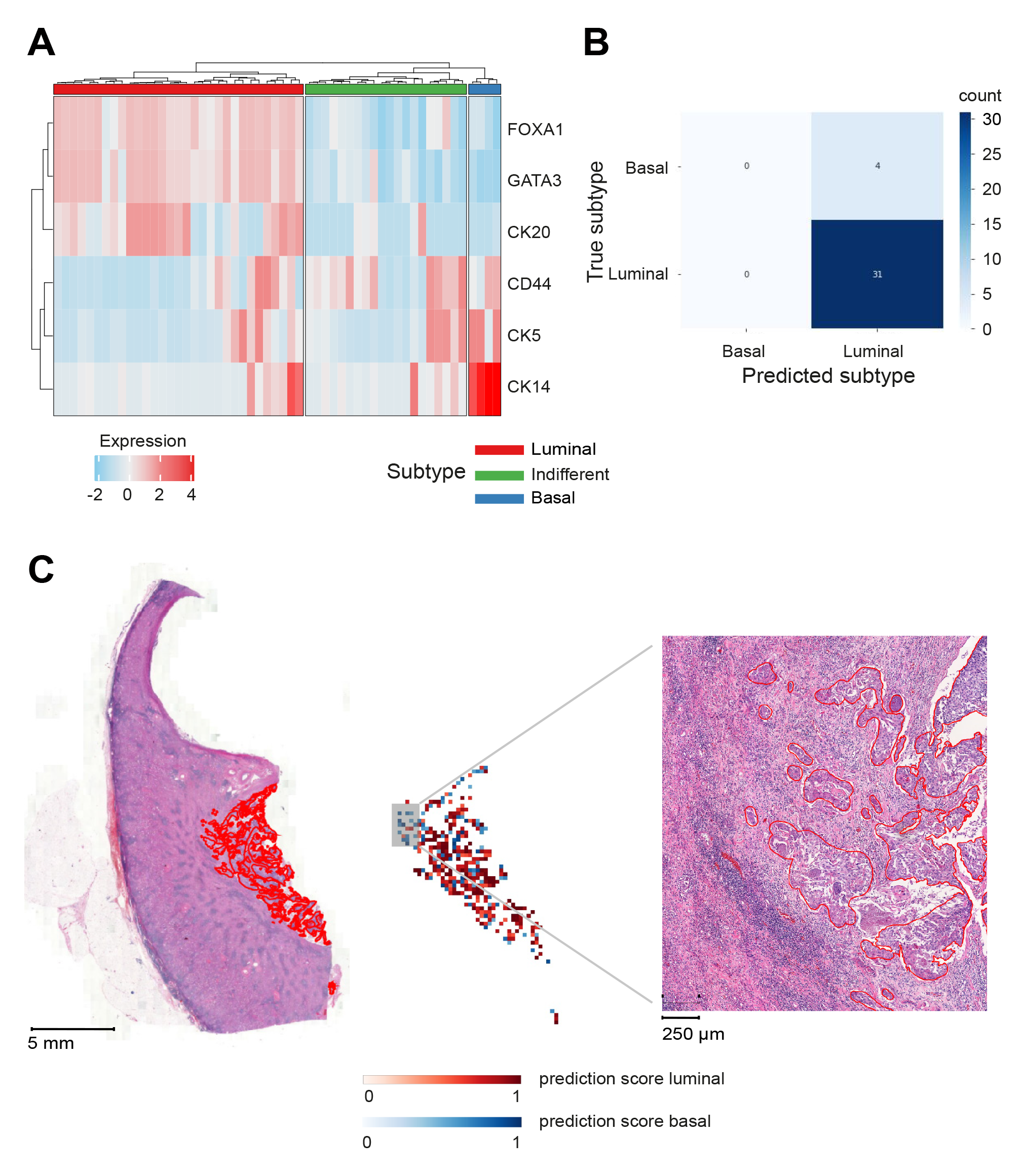
Validation of the deep-learning model on the Dutch cohort. (A) Heatmap visualization of the hierarchical clustering analysis performed on the expression of the three basal (CD44, CK5, CK14) and three luminal (FOXA1, GATA3, CK20) markers in the independent UTUC cohort from the Netherlands (N = 55 invasive samples). Heatmap colors represent markers expression (standardized H-Score; red: high expression, blue: low expression). The color ribbon at the top of the heatmap indicates the three protein-based subtypes: luminal (red), indifferent (green), basal (blue). (B) Confusion matrix with a summary of model’s generalization accuracy. (C) Selected basal slide predicted luminal by the model. From left to right: digitized whole-slide with annotated tumor areas (red); tile-level prediction map (red: luminal, blue: basal; intensity dependent on prediction score); zoomed area for a better visualization of the basal morphological features of the invasion front.

## Supplementary Tables

**Supplementary Table 1.** Association analysis of the identified subtypes with the main clinico-pathological variables. Estimates are given as median (minimum, maximum) or frequency (percentage) calculated on the total available samples. F: female; M: male. Kruskal-Wallis and Fisher’s exact tests were used respectively for continuous and categorical variables. Statistically significant p-values are highlighted in italic bold.

| Clinico-pathological variable | Luminal | Indifferent | Basal | p-value |
| --- | --- | --- | --- | --- |
| *Age at diagnosis (yrs), median (min-max)* | 72 (47-94) | 70 (48-85) | 77 (51-87) | ***0.02*** |
| *Gender, n (%)* |  |  |  | 0.07 |
| F | 24 (30) | 9 (22) | 19 (45.2) |  |
| M | 56 (70) | 32 (78) | 23 (54.8) |  |
| *Histological Grade (WHO 2016), n (%)* |  |  |  | 0.18 |
| low | 12 (15) | 7 (17.1) | 2 (4.8) |  |
| high | 68 (85) | 34 (82.9) | 40 (95.2) |  |
| *Primary Tumor, n (%)* |  |  |  | ***0.01*** |
| pT1 | 20 (25) | 10 (24.4) | 3 (7.1) |  |
| pT2 | 19 (23.8) | 4 (9.8) | 5 (11.9) |  |
| pT3 | 36 (45) | 21 (51.2) | 24 (57.1) |  |
| pT4 | 5 (6.2) | 6 (14.6) | 10 (23.8) |  |
| *Regional Lymph Nodes, n (%)* |  |  |  | 0.11 |
| N0 | 31 (77.5) | 11 (47.8) | 12 (57.1) |  |
| N1 | 4 (10) | 6 (26.1) | 6 (28.6) |  |
| N2 | 5 (12.5) | 6 (26.1) | 3 (14.3) |  |
| *Distant Metastasis, n (%)* |  |  |  | 0.57 |
| M0 | 36 (87.8) | 23 (95.8) | 20 (90.9) |  |
| M1 | 5 (12.2) | 1 (4.2) | 2 (9.1) |  |

**Supplementary Table 2.**
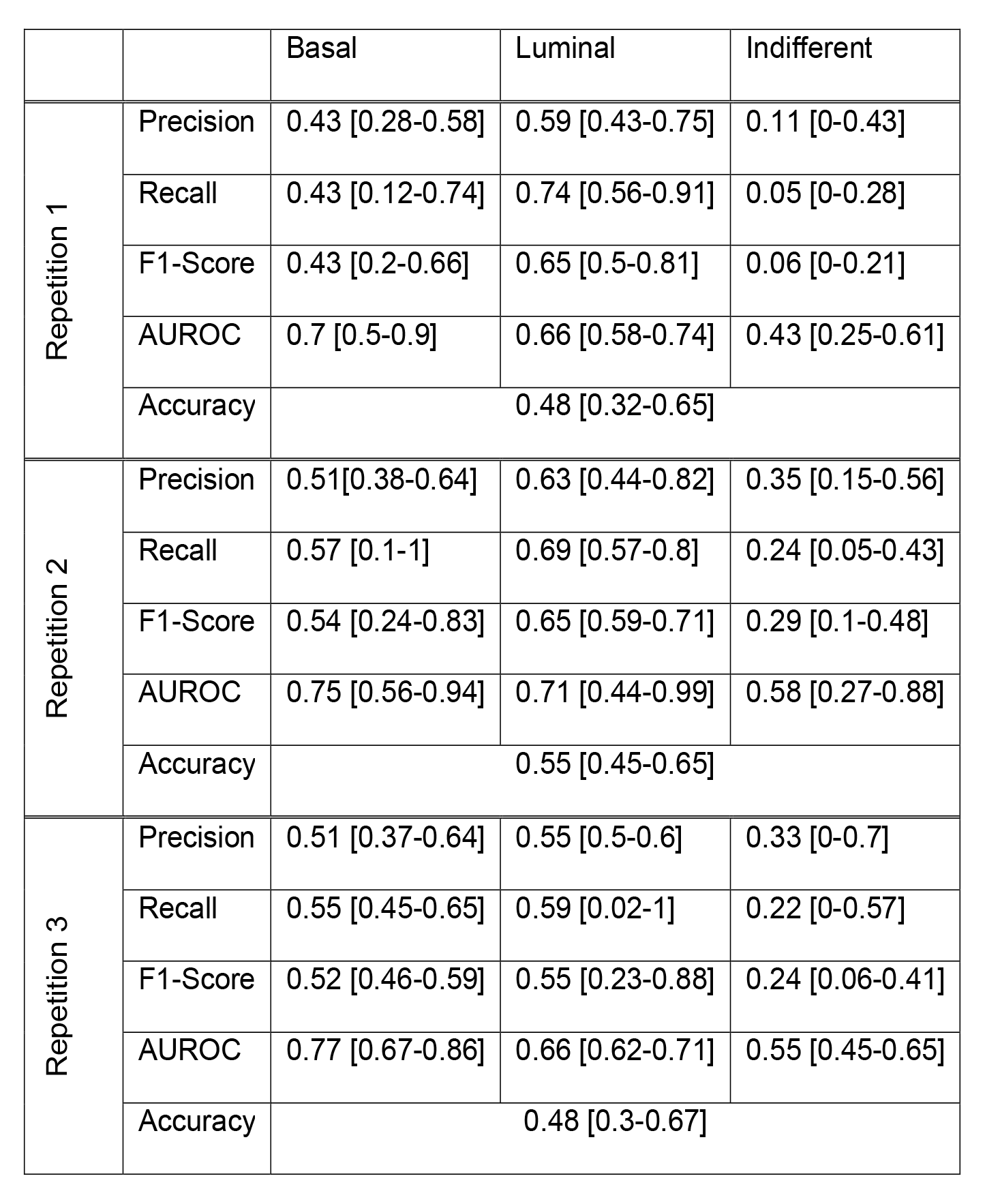
Performance metrics of the deep-learning classifier in the prediction of the luminal, basal, and indifferent protein-based subtypes for the three repetitions. For each repetition a measure of precision, recall, F1-Score, and AUROC is reported separately for the luminal, basal, and indifferent subtype together with the overall accuracy across the three groups. All performance metrics are indicated as mean [95% CI] across the hold-out folds. AUROC: area under the receiver operating characteristic; CI: confidence interval.

|  |  | Basal | Luminal | Indifferent |
| --- | --- | --- | --- | --- |
| Repetition 1 | Precision | 0.43 [0.28-0.58] | 0.59 [0.43-0.75] | 0.11 [0-0.43] |
|  | Recall | 0.43 [0.12-0.74] | 0.74 [0.56-0.91] | 0.05 [0-0.28] |
|  | F1-Score | 0.43 [0.2-0.66] | 0.65 [0.5-0.81] | 0.06 [0-0.21] |
|  | AUROC | 0.7 [0.5-0.9] | 0.66 [0.58-0.74] | 0.43 [0.25-0.61] |
|  | Accuracy | 0.48 [0.32-0.65] |  |  |
| Repetition 2 | Precision | 0.51 [0.38-0.64] | 0.63 [0.44-0.82] | 0.35 [0.15-0.56] |
|  | Recall | 0.57 [0.1-1] | 0.69 [0.57-0.8] | 0.24 [0.05-0.43] |
|  | F1-Score | 0.54 [0.24-0.83] | 0.65 [0.59-0.71] | 0.29 [0.1-0.48] |
|  | AUROC | 0.75 [0.56-0.94] | 0.71 [0.44-0.99] | 0.58 [0.27-0.88] |
|  | Accuracy | 0.55 [0.45-0.65] |  |  |
| Repetition 3 | Precision | 0.51 [0.37-0.64] | 0.55 [0.5-0.6] | 0.33 [0-0.7] |
|  | Recall | 0.55 [0.45-0.65] | 0.59 [0.02-1] | 0.22 [0-0.57] |
|  | F1-Score | 0.52 [0.46-0.59] | 0.55 [0.23-0.88] | 0.24 [0.06-0.41] |
|  | AUROC | 0.77 [0.67-0.86] | 0.66 [0.62-0.71] | 0.55 [0.45-0.65] |
|  | Accuracy | 0.48 [0.3-0.67] |  |  |

**Supplementary Table 3.**
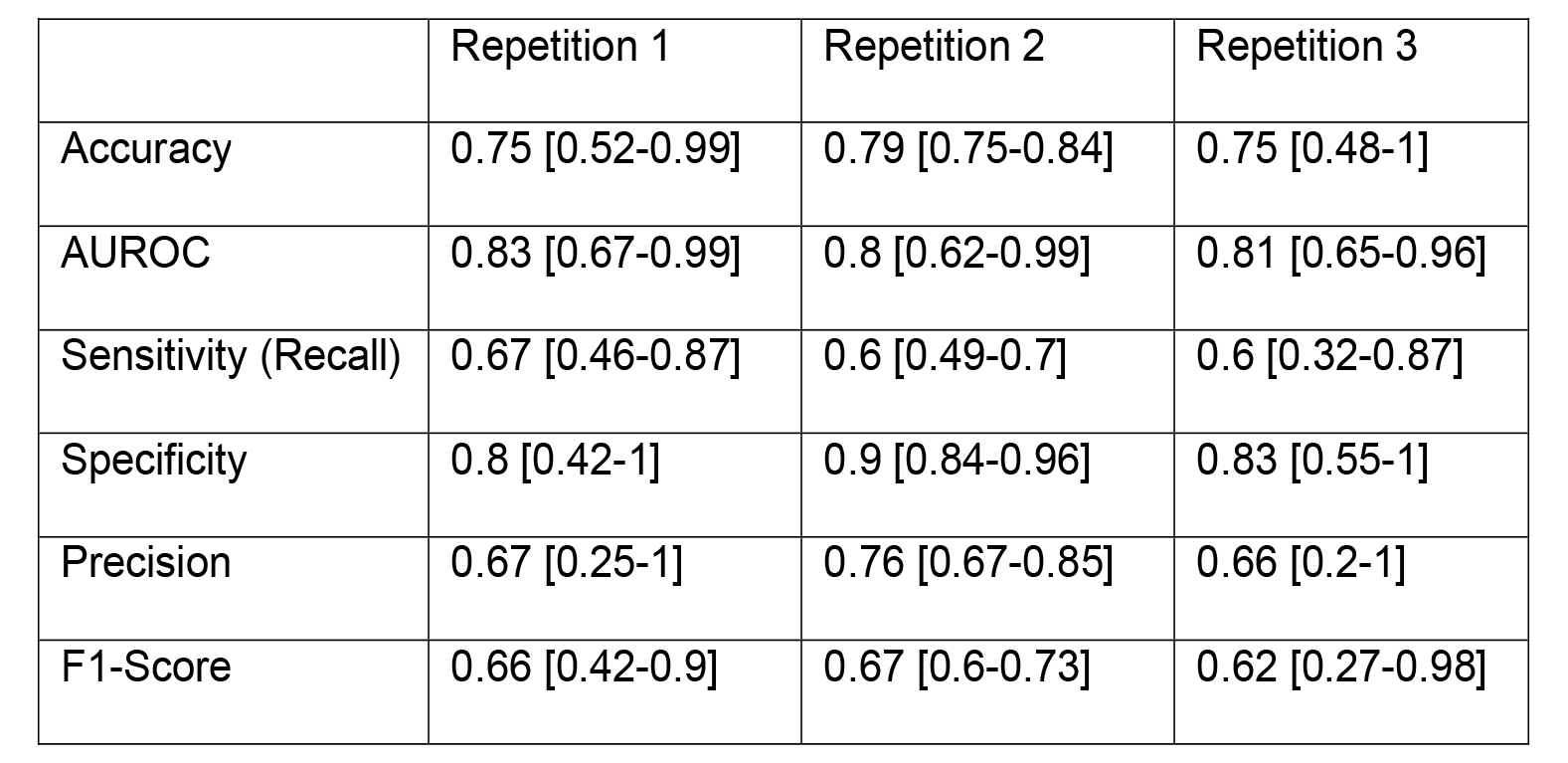
Performance metrics of the deep-learning classifier in the prediction of the luminal and basal protein-based subtypes for the three repetitions. For each repetition a measure of accuracy, AUC, sensitivity, specificity, precision, and F1-Score is reported for the basal subtype (here taken as the “positive” class) as mean [95% CI] across the hold-out folds. AUROC: area under the receiver operating characteristic; CI: confidence interval.

|  | Repetition 1 | Repetition 2 | Repetition 3 |
| --- | --- | --- | --- |
| Accuracy | 0.75 [0.52-0.99] | 0.79 [0.75-0.84] | 0.75 [0.48-1] |
| AUROC | 0.83 [0.67-0.99] | 0.8 [0.62-0.99] | 0.81 [0.65-0.96] |
| Sensitivity (Recall) | 0.67 [0.46-0.87] | 0.6 [0.49-0.7] | 0.6 [0.32-0.87] |
| Specificity | 0.8 [0.42-1] | 0.9 [0.84-0.96] | 0.83 [0.55-1] |
| Precision | 0.67 [0.25-1] | 0.76 [0.67-0.85] | 0.66 [0.2-1] |
| F1-Score | 0.66 [0.42-0.9] | 0.67 [0.6-0.73] | 0.62 [0.27-0.98] |

**Supplementary Table 4.** High-confidence predicted luminal and basal slides. The table lists all slides predicted luminal (sheet 1) or basal (sheet 2) with a prediction score higher or equal than 0.7. For each slide the true label (column “Subtype”), i.e., the label assigned by hierarchical clustering analysis, the prediction score for the basal (column “PredScore_Basal”) and luminal (column “PredScore_Luminal”) classes, the histological characterization (columns E-I), as well as the H-Score associated with the six markers panel, are reported. High-confidence predicted luminal and basal slides are sorted by descending values of PredScore_Luminal and PredScore_Basal respectively.

**Supplementary Table 5.** Low-confidence predicted slides. The table lists all slides for which the prediction score for the basal class was in the range [0.4-0.6]. For each slide the true label (column “Subtype”), i.e., the label assigned by hierarchical clustering analysis, the prediction score for the basal (column “PredScore_Basal”) and luminal (column “PredScore_Luminal”) classes, the histological characterization (columns E-I), as well as the H-Score associated with the six markers panel, are reported.

## Author contributions

**Conception and design:** Angeloni, Ferrazzi, Bahlinger

**Acquisition of data:** Angeloni, van Doeveren, Volland, Schmelmer, Stoehr, Geppert, Heers, Wach, Taubert, Sikic, Wullich, van Leenders, Eckstein, Boormans, Ferrazzi, Bahlinger

**Analysis and interpretation of data**: Angeloni, Lindner, Ferrazzi, Bahlinger

**Drafting of the manuscript:** Angeloni, Ferrazzi, Bahlinger

**Critical revision of the manuscript for important intellectual content:** All authors

**Statistical analysis:** Angeloni, Ferrazzi

**Obtaining funding:** Eckstein, Hartmann, Boormans, Ferrazzi

**Administrative, technical, or material support**: None

**Supervision:** Hartmann, Ferrazzi, Bahlinger

**Other:** Foersch (resources), Matek (resources), Zaburdaev (resources)

## Acknowledgments

We are grateful to Natascha Leicht, Christa Winkelmann, and Verena Popp for excellent technical assistance in the laboratory.

