## Supplementary Material for "A deep-learning workflow to predict upper tract urothelial cancer subtypes supporting the prioritization of patients for molecular testing"

### Supplementary Material and Methods for the paper *“A deep-learning workflow to predict upper tract urothelial cancer subtypes supporting the prioritization of patients for molecular testing”* by Angeloni et al.

### Tissue microarray analysis

Hematoxylin and Eosin (HE) staining, as well as tissue microarray (TMA) analysis, were performed for the two cohorts in the respective pathology centers. For each patient a total of four representative tissue cores (1 mm of diameter), two covering the tumor centrum and two covering the invasion front, were punched from the associated paraffin block and transferred to distinct recipient blocks using the TMA Grand Master (3DHistech, Hungary).

### Immunohistochemistry (IHC) analysis

All IHC analyses were performed at the Institute of Pathology, University Hospital Erlangen on a Ventana BenchMark Ultra (Ventana, USA) autostainer accredited by the German Accreditation Office (DAKKs) according to DIN EN ISO/IEC 17020. For protein-based subtyping, IHC staining with a six-marker panel consisting of three basal, i.e. CK14 (clone SP53, Cell Marque), CK5 (clone SP53, Cell Marque), CD44 (clone DF1485, Dako), and three luminal, i.e. CK20 (clone Ks 20.8, Dako), FOXA1 (polyclonal, Abcam), GATA3 (clone L50-823, DCS), protein markers was performed on 2-3 μm TMA sections from each block. The expression of these markers was histologically quantified (V.B. and P.V) using the histoscore (H-Score), which converts immunoreactivity into a semi-quantitative range [0-300] proportional to both staining intensity and percentage of positively stained cells. To validate the model’s predictions, immunohistochemical evaluations at the whole-slide level were performed using the same markers for selected cases.

PD-L1 expression on immune and tumor cells was assessed on TMAs using the Ventana PD-L1 assay (clone SP263) as previously described [1]. Quantification was performed by a pathologist (V.B.) relying on both immunoscore (IC) and combined positive score (CPS). The IC was calculated as the percentage of the area occupied PD-L1 positive immune cells relative to the total tumor area, whereas the CPS was calculated as the number of immune and tumor cells positive for PD-L1 out of the total number of tumor cells. Only samples with IC >= 5% or CPS >= 10 were considered positive for PD-L1 [2, 3].

### DNA isolation and *FGFR3* SNaPshot analysis

Tumor DNA was isolated using the Maxwell 16 LEV Blood DNA Kit (Promega, Mannheim, Germany) according to the manufacturer’s instructions as previously described [4]. *FGFR3* mutational analysis was performed using the SNaPshot method, as described in [5], which simultaneously detects nine hot spot mutations.

### Clustering-based protein subtypes identification and statistical analyses.

Clustering and statistical analyses were performed within the R environment v.4.0.3 [6].

To identify protein-based subtypes, the expression of each marker in each patient was taken equal to the median H-Score across the four TMA cores. Unsupervised hierarchical clustering was performed on the standardized markers expression (i.e. scaled to a mean of zero and a standard deviation of one) using Ward’s clustering method [7]. Ward’s algorithm was implemented relying on the R function *hclust* using as input the non-squared dissimilarity matrix computed through the Euclidean distance and ward.D2 as argument for the agglomeration method [8]. Heatmap visualization of the hierarchical clustering analysis was performed relying on the function *Heatmap* from the R-package ComplexHeatmap v.2.4.3.

Association analysis between categorical variables was performed using Fisher’s exact test. To compare the distribution of continuous variables, the Wilcoxon rank-sum test for independent samples (two groups) or the Kruskal-Wallis test (more than two) were used. Differences in the distribution of luminal and basal markers expression were evaluated using the one-tailed Wilcoxon signed-rank test for paired samples.

Analyses of overall survival (OS) and disease-specific survival (DSS) were performed using the Kaplan-Meier (K-M) estimator relying on the R-packages *survminer* v.0.4.9 and *survival* v.3.2.13. The statistical difference between survival curves was assessed through the log-rank test.

P-values < 0.05 were considered statistically significant.

### Slides digitization and WSIs annotation

Slides belonging to the two cohorts were digitized in the respective pathology centers using a Panoramic P250 scanner (3DHistech, Budapest, Hungary). Glass slides from the German cohort were scanned at 20-fold magnification with a resolution of 0.389 microns per pixel (mpp) whereas those from the Dutch cohort had three different resolution levels, i.e. 0.1214 mpp at 40-fold magnification, 0.2428 mpp at 20-fold magnification and 0.2484 mpp at 40-fold magnification. To facilitate the analysis and pre-processing of whole slide images (WSIs), digitized slides from the two cohorts were organized into two distinct QuPath (v.0.2.3) projects and stored as .qpproj files. Within each project, the tumor tissue belonging to the WSIs was manually annotated by a trained observer (M.A.) under the supervision of an expert uropathologist (V.B.). Manual annotation consisted in drawing a region of interest (ROI) around the tumor area and leaving out healthy tissue. Annotations were made at high levels of magnification using the brush and wand tools available in QuPath to exclude as much as possible non-tumor tissue including necrosis, bleeding/blood vessels, peri-tumoral lymphocytes and scanning artifacts (Supplementary Figure 1).

### WSIs pre-processing pipeline

Whole slide images (WSIs) pre-processing as well as deep learning analyses were performed in Python v.3.7.12 and run in a dedicated conda environment on a remote server based on Ubuntu’s 20.04.5 long-term support (LTS) operating system with NVIDIA Tesla V100-PCIE-32GB graphics processing unit (GPU).

An automated Python-based pipeline (<https://github.com/MiriamAng/TilGenPro>) was implemented for the pre-processing of WSIs. The pipeline only requires as input a QuPath project of annotated WSIs. First a groovy script is run to tessellate the identified tumor areas into smaller non-overlapping square patches of 512x512 pixel edge length. Subsequently, the generated tiles undergo a quality-filtering step. Namely, for a given WSI the median pixel intensity value across the RGB channels is calculated for each of the belonging tiles and a log_10_-transformed median pixel intensities distribution is obtained. A lower/upper percentile-based threshold can be set on the obtained distribution to filter-out tiles with a log_10_ median intensity lower or equal than the lower threshold (this corresponds to tiles characterized by darker regions) and/or greater or equal than the upper threshold (this corresponds to tiles with a high amount of white pixels). The pipeline was run using the 5^th^ percentile as lower threshold and the 90^th^ percentile as upper threshold. Finally, to reduce stain variation between training and test set, those tiles passing the quality-filtering step are stain-normalized according to the Macenko method [9] (Supplementary Figure 2). Macenko stain-normalization was implemented by assigning to the input parameters α and β the values of 1 and 0.15 respectively, as recommended by the authors [9], and using as reference H&E optical density (OD) matrix the one provided by Mitko Veta’s “Staining unmixing and normalization” code (<https://github.com/mitkovetta/staining-normalization>). For the Dutch cohort, non-overlapping square patches of 512x512 pixel edge length were generated taking as reference the resolution level of the German cohort (i.e. 0.389 mpp).

All tiles originating from a given slide inherited the corresponding patient-level label assigned by hierarchical clustering. A total of 341,906 (mean: 2098; range: 89-7515; 146,611 luminal; 109,681 basal; 85,614 indifferent) and 112,562 (mean: 2047; range: 58-4964; 57,312 luminal; 10,258 basal; 44,985 indifferent) tiles were generated for the German and the Dutch cohort respectively. To control, in training dataset composition, the over-representation of tiles from WSIs characterized by larger tumor area, thus maximizing the representativeness of tiles associated with smaller tumor areas, a maximum number of 2000 tiles was randomly subsampled from the WSIs of the German cohort used during training.

### Deep-learning algorithm and its validation

To predict protein-based subtypes in UTUC we relied on a transfer-learning approach by fine tuning a ResNet50 [10] initialized with weights pre-trained for the visual recognition challenge on the ImageNet database [11]. For model’s implementation the deep-learning library *fastai* v.1.0.61 [12], which is built on top of the open source PyTorch machine-learning framework [13], was employed. The pre-trained ResNet50 was retrieved from the *vision.learner* fastai module through the cnn_learner method. To adapt the pre-trained model to the specific classification task, model’s head was fine-tuned by setting a maximum number of 30 epochs using the 1cycle policy [14] with a maximum learning rate of 10^-5^ and a weight decay of 0.1. In addition to weight decay, other regularization techniques were implemented to avoid overfitting including data augmentation and early stopping. Data augmentation was performed relying on the fastai *get_transforms* function using the default random transformations (i.e. horizontal flipping, rotation, zooming, wrapping and lighting). To introduce rotational invariance also vertical flipping was adopted by setting to true the argument flip_vert. For early stopping implementation validation accuracy was chosen as quantity to be monitored throughout the whole training process. Notably, the fastai early stopping callback was implemented to terminate training after a patience time of three epochs with no improvement (min_delta = 0.01) of the monitored metric. The fastai save model callback was then used to save the model at the best epoch, i.e. the best model. During inference, a prediction value per subtype was assigned to each image tile. For each WSI, tile-level predictions were then averaged, class-wise, and the subtype predicted with the highest average prediction was assigned to the WSI.

A repeated three-fold cross-validation was used to estimate the model’s generalization accuracy and error. Here, to ensure independence between training and validation sets, the random splitting into the three folds was performed at patient level. Further, the splitting was performed in a stratified manner, i.e. preserving the percentage of samples for a given class within each partition. To this aim, we relied on the module *StratifiedKFold* from the scikit-learn package v.0.24.1 using a different value of the random state argument for each repetition and setting to true the shuffle parameter. At each round of the cross-validation, the deep-learning model was trained on two folds out of three and evaluated on the hold-out fold. Run times to fully train the DL model in a three-times repeated three-fold cross-validation were around 1 day.

To account for class imbalance, a tiles balancing procedure was implemented to equalize the number of tiles belonging to each class within the training set (Supplementary Figure 3). For a classification problem involving $N$ different classes, our proposed tiles balancing procedure relies on the following steps:

1. calculate the number of tiles to be balanced $t^{*}$ as the cumulative sum of the number of tiles belonging to the class with the lower number of associated WSIs (in case two or more classes have the same number of associated WSIs, start with the one with the lower number of tiles);
2. given $Nw_{C_{i}}$ the total number of WSIs belonging to each of the $N-1$ remaining classes ($i = 1,\ldots,N-1$), calculate the maximum number of tiles ${t^{*}}_{C_{i}}$to keep for each WSI belonging to class $i$ as ${t^{*}}_{C_{i}}=\frac{t^{*}}{Nw_{C_{i}}}$. If all the $Nw_{C_{i}}$WSIs have a number of tiles > ${t^{*}}_{C_{i}}$, then it would be sufficient to randomly choose a number equal to ${t^{*}}_{C_{i}}$ and the balancing procedure would be over.

If the condition in point 2 does not apply:

3. let $Nw_{C_{i}}^{lowEq}$ be the number of WSIs of class $i$ with a number of tiles $\leq$ ${t^{*}}_{C_{i}}$ and $Nw_{C_{i}}^{high}$ the number of WSIs of class $i$ with a number of tiles > ${t^{*}}_{C_{i}}$ . Given $t_{C_{i}}(j)$ the number of tiles belonging to the $j$-th WSI belonging to class $i$:
   a. keep all the tiles associated with the $Nw_{C_{i}}^{lowEq}$ WSIs and calculate the first contribution to the balancing as $t_{C_{i}}^{lowEq}=\sum_{j=1}^{Nw_{C_{i}}^{lowEq}} t_{C_{i}}(j)$
   b. calculate the new number of tiles ${t^{{high}^{*}}}_{C_{i}}$ to keep for the $Nw_{C_{i}}^{high}$ WSIs as ${t^{{high}^{*}}}_{C_{i}}=\frac{t^{*}- t_{C_{i}}^{lowEq}}{Nw_{C_{i}}^{high}}$ . If a WSI has a number of tiles <= ${t^{{high}^{*}}}_{C_{i}}$ keep them all (second contribution to the balancing for class $i$: $t_{C_{i}}^{high1}$), otherwise randomly choose a number equal to ${t^{{high}^{*}}}_{C_{i}}$ (third contribution to the balancing for class $i$: $t_{C_{i}}^{high2}$).

The balancing procedure will satisfy the following equation:

$$t^{*}\cong t_{C_{i}}^{lowEq}+t_{C_{i}}^{high1}+t_{C_{i}}^{high2}, for i = 1,\ldots,N-1$$

Performance metrics to evaluate model’s performance were calculated relying on the sklearn.metrics module. The area under the receiver operating curve (AUROC), accuracy, precision, recall, and F1-Score were assessed for each repetition, as mean across the three hold-out folds and 95% confidence interval (CI) relying on Student's t-distribution. Confusion matrices for a given repetition were instead obtained using the concatenated model’s predictions on the associated hold-out folds. As final model for the independent test cohort, a cross-validation ensemble was used. For each WSI of the test set, WSI-level predictions were obtained using each of the three models trained on the German cohort. Then, the final WSI-prediction was taken as the class (luminal/basal) with the highest average prediction value across the three models.
